# Religious affiliation and the risk of COVID 19 related mortality; a retrospective analysis of variation in pre and post lockdown risk by religious group in England and Wales

**DOI:** 10.1101/2020.10.01.20204495

**Authors:** Charlotte Hannah Gaughan, Daniel Ayoubkhani, Vahe Nafilyan, Peter Goldblatt, Chris White, Karen Tingay, Neil Bannister

## Abstract

**Background:** COVID 19 mortality risk is associated with demographic and behavioural factors; furthermore religious gatherings have been linked with the spread of COVID. We sought to understand the variation in the risk of COVID 19 related death across religious groups in the UK both before and after lockdown.

**Methods:** We conducted a retrospective cohort study of usual residents in England and Wales enumerated at the 2011 Census (n = 48,422,583), for risk of death involving COVID-19 using linked death certificates. Cox regression models were estimated to compare risks between religious groups. Time dependent religion coefficients were added to the model allowing hazard ratios (HRs) pre and post lockdown period to be estimated separately.

**Results:** Compared to Christians all religious groups had an elevated risk of death involving COVID-19; the largest age adjusted HRs were for Muslim and Jewish males at 2.5 (95% confidence interval 2.3-2.7) and 2.1 (1.9-2.5), respectively. The corresponding HRs for Muslim and Jewish females were 1.9 (1.7-2.1) and 1.5 (1.7-2.1). The difference in risk between groups contracted after lockdown. Those who affiliated with no religion had the lowest risk of COVID 19 related death before and after lockdown.

**Conclusion:** The majority of the variation in COVID 19 mortality risk was explained by controlling for socio demographic and geographic determinants; however, Jews remained at a higher risk of death compared to all other groups. Lockdown measures were associated with reduced differences in COVID 19 mortality rates between religious groups, further research is required to understand the causal mechanisms.

## Introduction

The probability to become infected and subsequently die from Coronavirus disease 2019 (COVID 19) has been shown to vary depending on a variety of factors including socio economic determinants and behavioural factors [1]. Despite concerns expressed by the World Health Organisation (WHO) that religious practises can contribute to the spread of COVID 19 [2], little is known about the differing risk of mortality to religious groups. For example, extended transmission during communal religious prayers and large attendance at religious gatherings and festivals may be factors in community transmission [3]. Cultural factors, such as contact with large extended families and strong community links, are also considered likely factors in the spread amongst religious communities [2]. Furthermore, several studies have shown that outbreaks worldwide have been traced to centres of worship and religious ceremonies [3,4].

Religious gatherings were prohibited as part of a sweep of measures introduced into UK law on 23 March 2020 with the aim of preventing the spread of COVID 19. In this paper, we investigate whether the risk of COVID-19 mortality differs between religious groups in England and Wales. Around 67% of the UK population state that they identify with a religion [5]. Whilst affiliation to a religious group does not necessarily equate to religious practice (75% of Sikhs attend regular practises whilst only 29% of Christians do) [5], quantifying the extent to which risk differs between religious groups overall is valuable in assessing inequalities in COVID-19 mortality.

Using data from the 2011 Census of England and Wales and linked death registrations, we estimated the age-adjusted risk of dying with COVID 19 for each religious group; we then use socio demographic information, indicators of occupational exposure and geographical measures to adjust for factors related to both the spread of the virus in the general population and the potential increased risk of death following infection as a result of inequality.

We make two contributions to the literature on disparities in COVID-19 outcomes. First, whilst evidence suggests that religious services may spread infection [2, 3], to the best of our knowledge, no study has specifically examined the mortality risk of different religious groups, and specifically after adjusting for socio demographic, occupational and geographical determinants. Second, this study examines the association between state mandated prohibition of religious services and COVID 19 mortality risk across different religious groups.

## Methods

### Study Population

The study population included all usual residents enumerated in private households in England and Wales at the 2011 Census who were not known to have died before 2 March 2020 (n = 48,422,583) according to death certificates linked to the Census on NHS Number.

We analysed all COVID 19 deaths (n = 36,726) and a weighted 1% sample of other individuals in the study population. The sample was further down-weighted to account for possible differential propensities to emigrate between Census date (27 March 2011) and 1 March 2020 across religions; these weights were calculated based on data from the NHS Patient Register and the outflows of the International Passenger Survey and the ONS Longitudinal Survey. The datasets were deterministically linked using NHS number. In addition, those enumerated in March 2011 answering the “Intention to Stay” for less than six months were excluded from the analyses because of their high propensity to have left the UK before the analysis period under investigation. Since this study was a follow up of the 2011 census we were unable to include people who were born or migrated into England and Wales since March 2011.

### Outcome Variable

Deaths occurring between 2 March 2020 and 15 May 2020, and registered by 29 May 2020, were classified as being related to COVID 19 if ICD-10 codes U07.1 (virus identified) or U07.2 (virus not identified) were mentioned on the death certificate, either as the cause of death or as a contributing factor.

### Exposure Variable

Self-reported religious affiliation from the 2011 Census was used to assign individuals to one of nine groups: No religion, Christian, Buddhist, Hindu, Jewish, Muslim, Sikh, Any other religion, or Religion not stated.

### Covariates

All of the covariates described below and included in the model were taken from the 2011 Census. Age was included as a second-order polynomial to account for the non-linear relationship between age and risk of death. The other covariates were grouped together to control for the effects of geography, socio economic status, household characteristics, occupation exposure and ethnicity.

Place of usual residence in 2011 was used for both the region and population density variables (population density itself was based on the 2018 mid-year estimate).[8] Population density was included in the model as a second-order polynomial, with an interaction term included to allow the slope to vary beyond the 99^th^ percentile of the distribution, which accounts for extreme values. Socio economic status (NS-SEC), education, and deprivation [9,10] were grouped together to capture socio economic determinants of health. Self-reported health was a measure ranging from very good to very bad in 2011. Household composition accounted for the number of people in a household, the presence of a multi-generational household, and main language spoken in the household. Variables indicating whether an individual was a keyworker, or if there was a keyworker in the household, were derived according to previous ONS methodology [11] Finally, owing to evidence that people of non-white ethnicity have been disproportionately affected by COVID in the UK [1], a binary white / non-white indicator variable was included in the model to refine any residual religion specific variation to be quantified.

### Statistical Analysis

We quantified the absolute risk of COVID 19 related death across religious groups using age-standardised mortality rates (ASMRs) [6], where the mortality rates were standardised according to the overall age-sex distribution in the study population.

We used Cox regression models to estimate hazard ratios (HRs) for COVID 19 related death across religious groups, using Christian (the largest group) as the reference category. Separate models were estimated for males and females. Descriptions of the models can be found in the supplementary materials. Follow-up time ran from 2 March 2020 to 15 May 2020, and individual’s contribution to the risk of death was censored if they did not die with COVID 19 during this period. To assess the total association between risk profile of religious groups and lockdown measures, HRs were estimated from time-dependent coefficients by dividing follow-up time into pre and post lockdown spells [7]. The 10^th^ April, two weeks after lockdown measures came into force on 23 March 2020, was used as the cut-off date to allow a lag of 14 days between pre lockdown COVID 19 infection and subsequent death.

## Results

In our study population the mean age was 47 years (minimum 9 years, maximum 110 years). Over the outcome period, mean follow-up time was 73.9 days (standard deviation 2.2 days). For both males and females, Jews, Muslims, Hindus, Sikhs and Buddhists experienced greater age-adjusted rates of COVID 19 related mortality than Christians, while the rates were lower still among those with no religion (table 1).

### Absolute risk of COVID 19 related death by religion

**Table 1.** Age standardised mortality rates (and 95% confidence intervals) for COVID 19 related death, stratified by religious affiliation and sex.

|  | **Males**<br>**(n = )** | **Females**<br>**(n = )** |
| --- | --- | --- |
| **No religion**<br>**(n = )** | 80.3 (76.9-83.8) | 47.2 (44.1-50.3) |
| **Christian**<br>**(n = )** | 92.6 (90.9-93.8) | 54.6 (53.3-55.2) |
| **Buddhist**<br>**(n = )** | 113.5 (77.3-140.2) | 57.4 (37.8-79.2) |
| **Hindu**<br>**(n = )** | 154.8 (138.0-171.5) | 93.3 (78.8-103.0) |
| **Jewish**<br>**(n = )** | 187.9 (164.6-210.8) | 94.3 (75.6-104.0) |
| **Muslim**<br>**(n = )** | 198.9 (181.4-209.4) | 98.2 (86.3-105.7) |
| **Sikh**<br>**(n = )** | 128.6 (108.4-149.7) | 69.4 (54.5-83.0) |
| **Other Religion**<br>**(n = )** | 87.1 (64.3-115.2) | 45.1 (31.7-62.0) |
| **Not stated**<br>**(n = )** | 82.3 (78.0-86.6) | 48.9 (45.9-51.8) |

### Mediators of the relationship between religion and COVID 19 mortality

After adjusting for age, the differences in COVID 19 mortality rates between religious groups were larger for males than females (figure 1, and supplementary tables 1 and 2). Controlling for region and population density notably reduced the elevation in risk compared to the Christian population for all religious groups. The hazard ratio is reduced for Buddhist males to 1.0 (95% confidence interval [CI]:0.8-1.3) and females 0.8 (0.6-1.1), Hindu males 1.3 (1.2-1.5) and females 1.3 (1.1-1.5), Jewish males 1.5 (1.4-1.7) and females 1.1 (1.0-1.3), Muslim Males 1.7 (1.6 - 1.8) and Muslim females 1.3 (1.2-1.5), Sikh males 1.2 (1.0-1.4) and Sikh females 1.0 (0.8-1.2).

**Figure 1:**
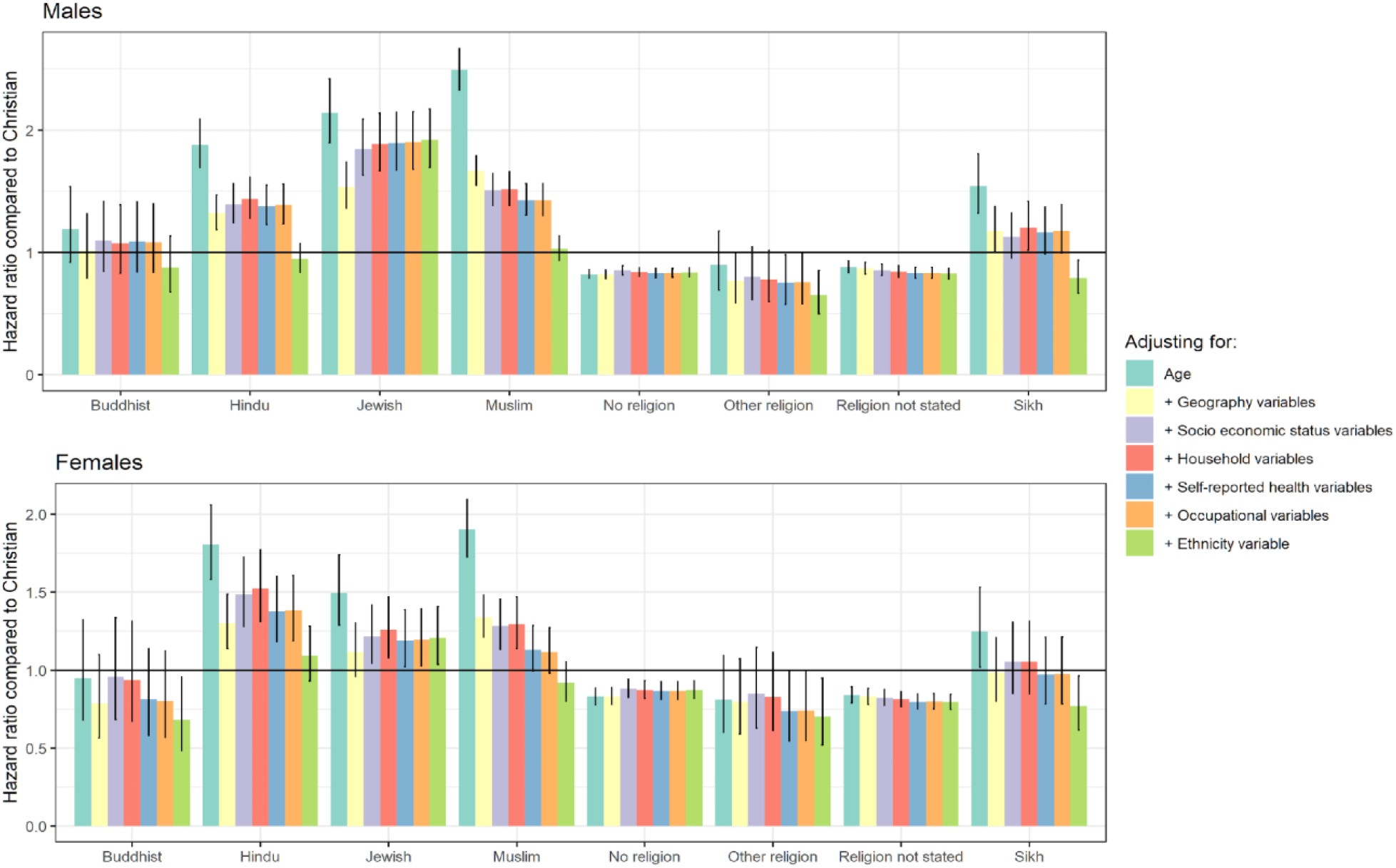
Hazard ratios for COVID 19 related mortality for religious groups compared to Christians, stratified by sex.

The inclusion of socio economic determinants shows some further reduction of risk for Muslims males HR 1.4 (1.3-1.6) and females 1.1 (1-1.3) however for other religious groups the inclusion of further covariates are negligible until the inclusion of ethnic group which reduces the risk for all main religions, except Jews for whom the hazard ratios increase to 1.9 (1.7-2.2) and 1.2 (1.0,1.4), respectively.

The impact on the HRs of adjusting for additional covariates was generally more subtle, at least until ethnicity was included in the models, which markedly reduced the HRs for Buddhists, Hindus, Muslims, and Sikhs. In the fully adjusted models, the HRs were close to unity for most religious groups, and mortality risk was in fact lower for some groups (Sikhs, female Buddhists, and those of any other religion, no religion, or unknown religious affiliation) compared to Christians. However, mortality risk remained elevated for Jewish males and females, with fully adjusted hazard ratios of 1.9 (1.7-2.2) and 1.2 (1.0-1.4), respectively.

### The evolution of the total association between religion and COVID 19 over the course of the pandemic

In order to assess the change in relative risk overtime, we have calculated Shoenfeld residuals, these represent the difference between the observed covariate and the expected value given the risk set at that time; we present the smoothed residuals which provide an estimate of how the residuals for each religious group evolve over the follow-up period.

The Schoenfeld residuals from the fitted Cox models suggest non-constant HRs for the Jewish, Muslim, Hindu and Sikh groups (supplementary figures 1 and 2). The elevation in mortality risk relative to the Christian population decreased over the course of the pandemic for these religious groups, most notably amongst Jews (figure 2).

**Figure 2:**
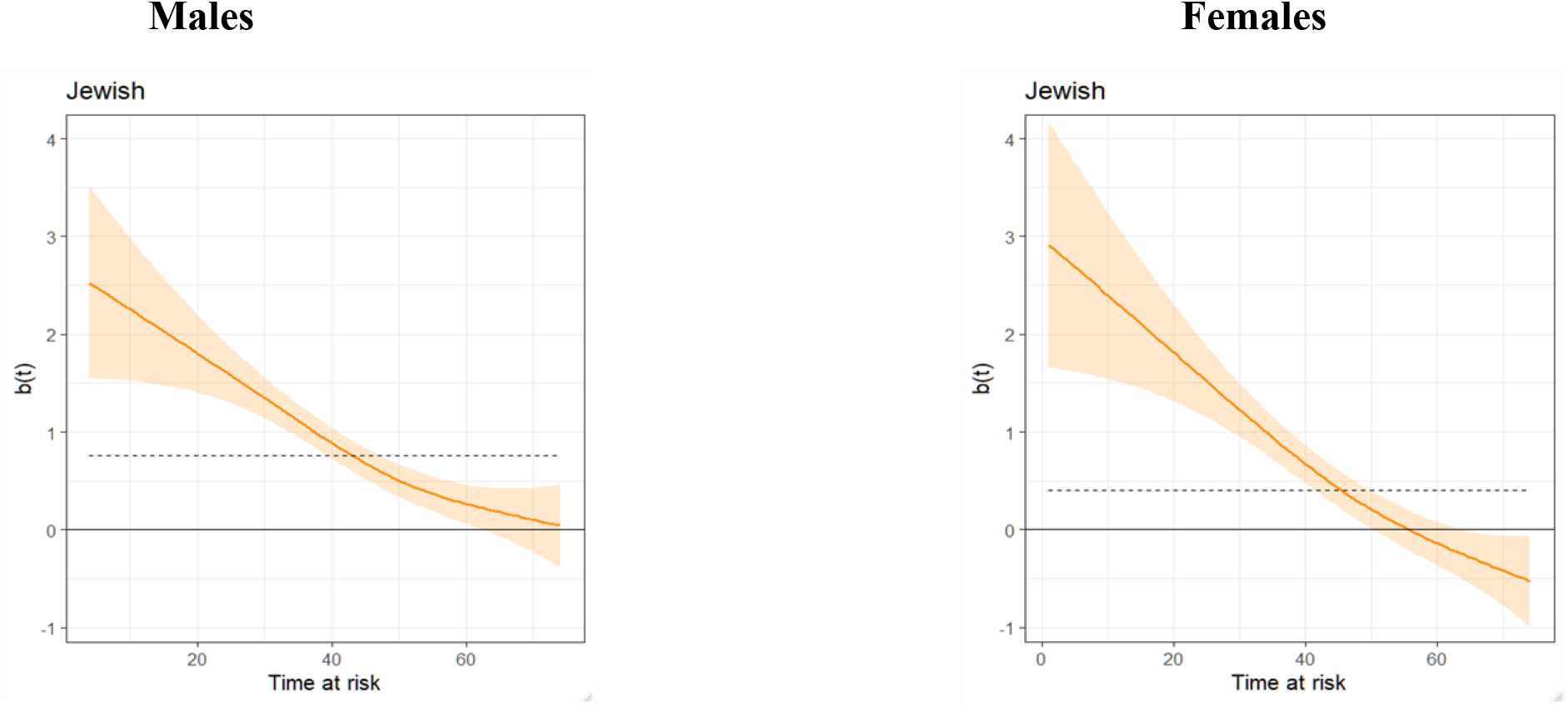
Smoothed Schoenfeld residuals from age adjusted Cox regression models for Jewish males and females Time at risk starts on 2 March 2020. b(t) represents the estimated time-varying model coefficient (the natural logarithm of the hazard ratio)

Figure 3 shows the age adjusted model to identify total association between groups as opposed to direct association. After stratifying the Cox model coefficients on pre and post lockdown periods, COVID 19 mortality risk before lockdown was considerably greater for Muslims (Male 4.0 (95% confidence interval [CI]: 3.6-4.4), Female 3.6 (3.-,4.2), Jews (Male 3.5 (2.9-4.2), Female 3.0 (2.4-3.8)) and Hindus (Male HR = 2.6 (2.2-3.0), Female HR = 3.5 (2.9-4.4)) compared with Christians.

**Figure 3:**
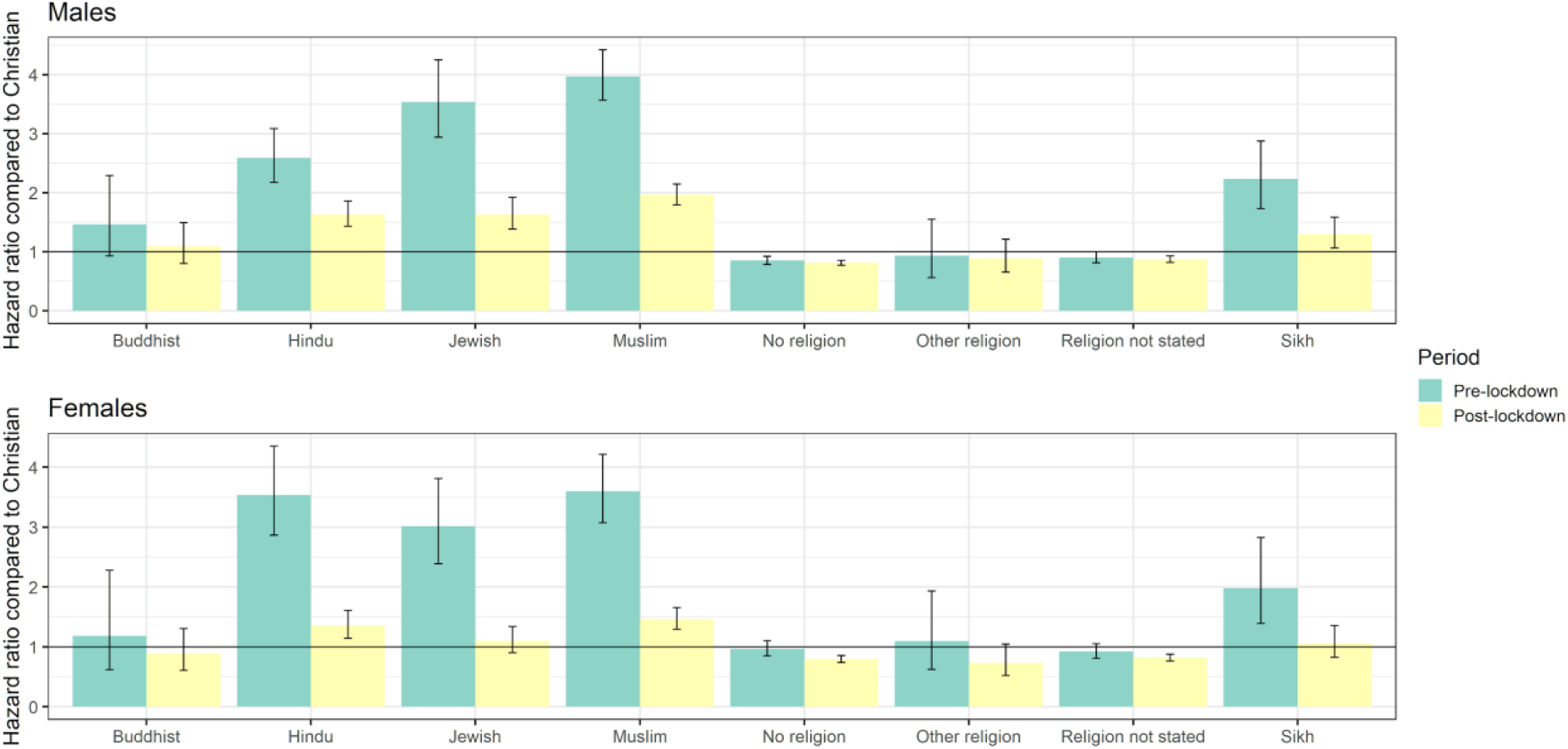
Pre and post lockdown, age adjusted hazard ratios of COVID 19 related mortality for religious groups compared to Christians, stratified by sex.

COVID 19 mortality risk before lockdown was elevated for Hindus, Jews, Muslims and Sikhs compared with Christians. The HRs for these four religious groups were notably reduced following lockdown for males, though all four remained greater than unity, at 1.6 (1.4-1.9), 1.6 (1.4-1.9), 2.0 (1.8-2.1) and 1.3 (1.1-1.6), respectively. Heterogeneity in COVID 19 mortality risk was reduced following lockdown to a greater extent for females than males, though the HRs remained elevated for Hindus and Muslims, at 1.4 (1.1-1.6) and 1.5 (1.3-1.7), respectively.

For all non Christian groups the national lockdown was associated with reduced mortality risk compared with the Christian group.

## Discussion

We analysed COVID 19 deaths between 2 March 2020 and 15 May 2020 linked to Census data to understand the risks to religious groups. The age adjusted rates show an elevated risk of COVID 19 mortality for Hindus, Jews, Muslims, Sikhs and Buddhists compared to the Christian population. Those affiliating with no religion were at a lower risk than their Christian counterparts. Compared with the age adjusted results, the estimated hazard ratios for religious groups were reduced when covariates were included in the models, indicating that geographical and socio-demographic factors to some extent mediate the relationship between religion and COVID 19 mortality. The HRs for individuals of no religion remained relatively constant as covariates were added to the models.

Including the pre and post lockdown risk models give us an indication of the risk to non Christian religious groups of the uncontrolled spread of COVID, and how the risk to religious groups changed as the result of government measures. Whilst the risk both pre and post lockdown is highest for Muslims, Jews and Hindus, the variation in the risk between religious groups is reduced. It is notable the risk to Jewish men and women was particularly high in the pre lockdown period.

Our results confirm that COVID 19 mortality risk for each non Christian group is in general higher for males than females, and where heterogeneity in risk is observed between religious groups, the elevation in risk compared to the Christian group is generally greater for males than females. We observe a large and unexplained increased risk for Jewish males; after controlling for geographic factors, the relative risk increased slightly as additional factors were included in the model. Jews had a raised risk despite being relatively advantaged in terms of the risk factors that contributed to higher mortality in predominantly non-white religious groups.

We include ethnicity (white, non-white) as the final covariate in the model, however as demonstrated in supplementary table 4 some religions (Buddhist, Hindu, Muslim and Sikh) are largely focused on non-white groups and therefore there is considerable confounding between their religion and non-white risk factors.

Our findings suggest that behavioural changes as a result of the lockdown and intervention measures operated to reduce the risk for religious groups, which may be a consequence of restrictions on congregating in places of worship. However, it is not possible from this analysis to confirm whether the reduction in risk to religious groups comes as a result of preventing other activities (e.g. prohibiting households from mixing, ordering pubs to close etc.) as opposed to specifically the banning of religious gatherings.

Whilst behaviours relating to religious practises could be responsible for higher infection rates leading to higher mortality rates, is not clear that all the residual risk from religion is a consequence of behaviour. For example, the impact of racial prejudice and self-reported racism has been shown to increase the risk of stress in ethnic and religious communities resulting in higher prevalence of illness including the impacts anticipatory stress [12,13,14]. In light of the extant levels of religious prejudice in UK society [15], it is highly possible this is experienced by religious communities and therefore it is possible that part of the increased risk seen in both the age adjusted and fully adjusted models are a result of a stress induced conditions resulting from religious prejudices.

A limitation of our study that the data were taken from the 2011 Census. Whilst using variables from 2011 for a population at risk of COVID 19 related death in 2020 is not ideal, it seems unlikely that religious affiliation will change over this period for the majority of people, although the extent to which they practice may do. Similarly, whilst the socio demographic factors included in our models may of course evolve over time for the individual, a significant shift in the profile of a religious group over the last nine years seems unlikely; for example, analysis of data from the Annual Population Survey (APS) [16] showed that between 2012 and 2019, the socio economic make-up of religious groups has generally not changed (see supplementary table 6). However, key worker status is likely to be out of date for a proportion of the population. It could also be the case that religious communities centre around religious buildings and not move over time so it is likely the geographical measures are accurate for people who most strongly practice their religion; furthermore, population density is reflective of the situation in 2018. The main problem is defining what religious affiliation at the 2011 Census is in practise measuring; in an ideal world the effect of religion would be specific to religious practices, however we are unable to observe the extent of religious practises in this study.

Whilst we may not have adjusted for all the socio-demographic determinants of religion, and therefore part of the increased risk of religion could be due to unobserved factors such as health status at the time of infection, the benefits of including further covariates are debatable. The covariates included cover a wide range of socio demographic variables that contribute to raised mortality involving COVID-19 in non-Christian religious groups. The residual risk after adjusting for these factors provides an indication of the specific effect of religious affiliation.

### What is already known on this subject

Certain religious services and practises have been shown to be catalysts for outbreaks of COVID 19.

The risk of COVID 19 related death is correlated with measures of inequality and ethnicity.

### What this study adds

People in England and Wales identifying with major religious groups experience a higher age-adjusted rate of COVID 19 related mortality than those of no religion

Geographic and socio-demographic factors mediate the relationship between religious group and risk of COVID 19 mortality, however there is an unexplained residual elevation in risk for Jews.

The introduction of lockdown measures was associated with reduced heterogeneity in COVID 19 mortality risk between religious groups.

## Data Availability

Data are presented in the supplementary material. Under the provisions of the Statistics and Registration Service Act 2007, the linked 2011 Census data used in this study are not permitted to be shared.

## Contributors

CG wrote the first draft of the manuscript. VN CW CG and DA designed the analysis. CG DA and VN conducted statistical modelling. All authors edited and reviewed manuscript.

## Acknowledgements

The authors would like to thank Jessica Walkeden, Alex Cooke, Louisa Blackwell, Rachel Shipsey and Merilynn Pratt at the Office for National Statistics who prepared the linked datasets and assisted with the statistical analysis.

## Patient consent for publication

Not required.

## Provenance and peer review

Not commissioned; externally peer reviewed.

