## Supplementary tables for "Religious affiliation and the risk of COVID 19 related mortality; a retrospective analysis of variation in pre and post lockdown risk by religious group in England and Wales"

*Supplementary Table 1. Hazard ratios (and 95% confidence intervals) for COVID-19 related death for religious groups compared to the Christian population, males*

| | Model 1 | Model 2 | Model 3 | Model 4 |
|---|---|---|---|---|
| | +Age | +Geography Variables | +Socio economic status variables | +Household variables |
| **No religion** | 0.82 (0.79-0.86) | 0.82 (0.78-0.86) | 0.85 (0.82-0.89) | 0.84 (0.80-0.88) |
| **Buddhist** | 1.19 (0.92-1.54) | 1.02 (0.79-1.31) | 1.09 (0.85-1.42) | 1.07 (0.83-1.39) |
| **Hindu** | 1.88 (1.69-2.09) | 1.32 (1.19-1.47) | 1.39 (1.24-1.56) | 1.44 (1.28-1.61) |
| **Jewish** | 2.14 (1.89-2.42) | 1.54 (1.36-1.74) | 1.84 (1.63-2.09) | 1.89 (1.67-2.14) |
| **Muslim** | 2.49 (2.32-2.67) | 1.67 (1.55-1.79) | 1.51 (1.38-1.64) | 1.52 (1.39-1.66) |
| **Sikh** | 1.54 (1.32-1.81) | 1.18 (1.00-1.38) | 1.13 (0.95-1.33) | 1.20 (1.02-1.42) |
| **Other** | 0.90 (0.69-1.17) | 0.77 (0.59-1.00) | 0.80 (0.61-1.04) | 0.78 (0.60-1.02) |
| **Religion not stated** | 0.88 (0.83-0.93) | 0.87 (0.82-0.92) | 0.86 (0.81-0.90) | 0.84 (0.80-0.89) |

| | Model 5 | Model 6 | Model 7 |
|---|---|---|---|
| | Self-reported health variables | Occupational Exposure variables | Ethnicity |
| **No religion** | 0.83 (0.80-0.87) | 0.83 (0.80-0.87) | 0.84 (0.80-0.87) |
| **Buddhist** | 1.09 (0.84-1.41) | 1.08 (0.84-1.40) | 0.88 (0.68-1.14) |
| **Hindu** | 1.38 (1.23-1.55) | 1.39 (1.24-1.56) | 0.95 (0.84-1.07) |
| **Jewish** | 1.89 (1.67-2.14) | 1.90 (1.68-2.15) | 1.92 (1.69-2.17) |
| **Muslim** | 1.43 (1.30-1.56) | 1.43 (1.30-1.56) | 1.03 (0.94-1.13) |
| **Sikh** | 1.16 (0.99-1.37) | 1.18 (1.00-1.39) | 0.79 (0.67-0.94) |
| **Other** | 0.75 (0.58-0.98) | 0.76 (0.58-0.99) | 0.65 (0.50-0.85) |
| **Religion not stated** | 0.83 (0.79-0.88) | 0.83 (0.79-0.88) | 0.83 (0.78-0.87) |

*Supplementary Table 2. Hazard ratios (and 95% confidence intervals) for COVID-19 related death for religious groups compared to the Christian population, females*

| | Model 1 | Model 2 | Model 3 | Model 4 |
|---|---|---|---|---|
| | +Age | +Geography Variables | +Socio economic status variables | +Household variables |
| **No religion** | 0.83 (0.78-0.89) | 0.83 (0.78-0.89) | 0.88 (0.82-0.94) | 0.87 (0.82-0.93) |
| **Buddhist** | 0.95 (0.68-1.32) | 0.79 (0.57-1.10) | 0.96 (0.68-1.34) | 0.94 (0.67-1.31) |
| **Hindu** | 1.81 (1.58-2.06) | 1.30 (1.14-1.49) | 1.48 (1.28-1.72) | 1.52 (1.31-1.77) |
| **Jewish** | 1.50 (1.29-1.74) | 1.12 (0.96-1.30) | 1.22 (1.04-1.42) | 1.26 (1.08-1.47) |
| **Muslim** | 1.90 (1.72-2.10) | 1.34 (1.21-1.48) | 1.28 (1.13-1.46) | 1.29 (1.14-1.47) |
| **Sikh** | 1.25 (1.02-1.53) | 0.98 (0.80-1.21) | 1.05 (0.85-1.31) | 1.05 (0.85-1.31) |
| **Other** | 0.81 (0.60-1.09) | 0.80 (0.59-1.07) | 0.85 (0.63-1.15) | 0.83 (0.61-1.12) |

| | | | | |
|---|---|---|---|---|
| **Religion not stated** | 0.84 (0.79-0.89) | 0.83 (0.78-0.88) | 0.82 (0.77-0.88) | 0.81 (0.76-0.86) |

| | **Model 5**<br>**+Self-reported health variables** | **Model 6**<br>**+Occupational Exposure variables** | **Model 7**<br>**+Ethnicity** |
|---|---|---|---|
| **No religion** | 0.87 (0.81-0.93) | 0.87 (0.81-0.93) | 0.87 (0.82-0.93) |
| **Buddhist** | 0.81 (0.58-1.14) | 0.80 (0.57-1.12) | 0.68 (0.48-0.96) |
| **Hindu** | 1.38 (1.18-1.60) | 1.38 (1.19-1.61) | 1.09 (0.93-1.28) |
| **Jewish** | 1.19 (1.02-1.39) | 1.20 (1.03-1.39) | 1.21 (1.04-1.41) |
| **Muslim** | 1.13 (0.99-1.29) | 1.12 (0.98-1.27) | 0.92 (0.80-1.05) |
| **Sikh** | 0.97 (0.78-1.21) | 0.97 (0.78-1.21) | 0.77 (0.61-0.96) |
| **Other** | 0.74 (0.54-0.99) | 0.74 (0.55-1.00) | 0.70 (0.52-0.95) |
| **Religion not stated** | 0.80 (0.75-0.85) | 0.80 (0.75-0.85) | 0.79 (0.75-0.85) |

*Supplementary figure 1. Male Schoenfeld residuals from Age adjusted Cox Regression model*

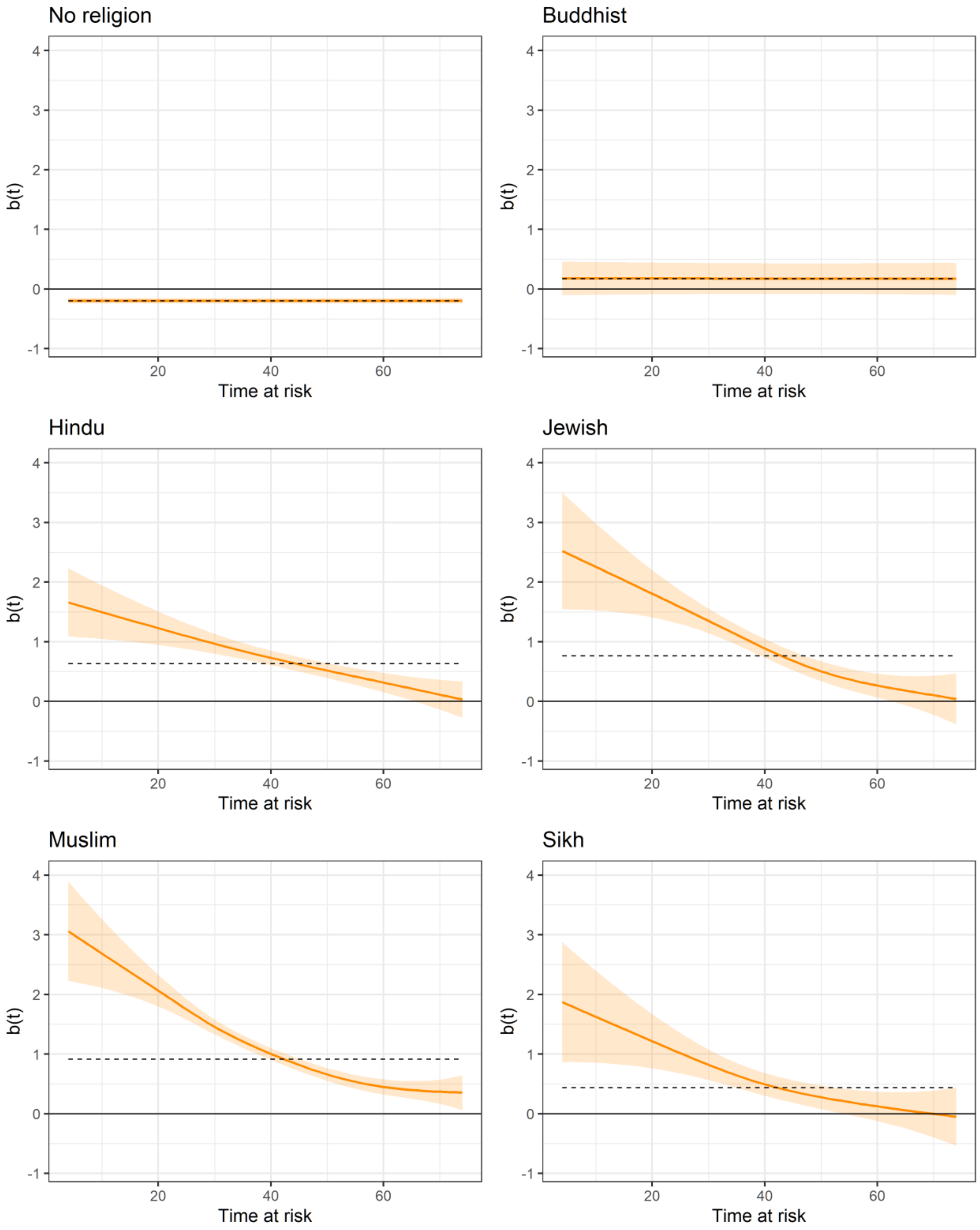

*Supplementary figure 2. Female Schoenfeld residuals from Age adjusted Cox Regression model*

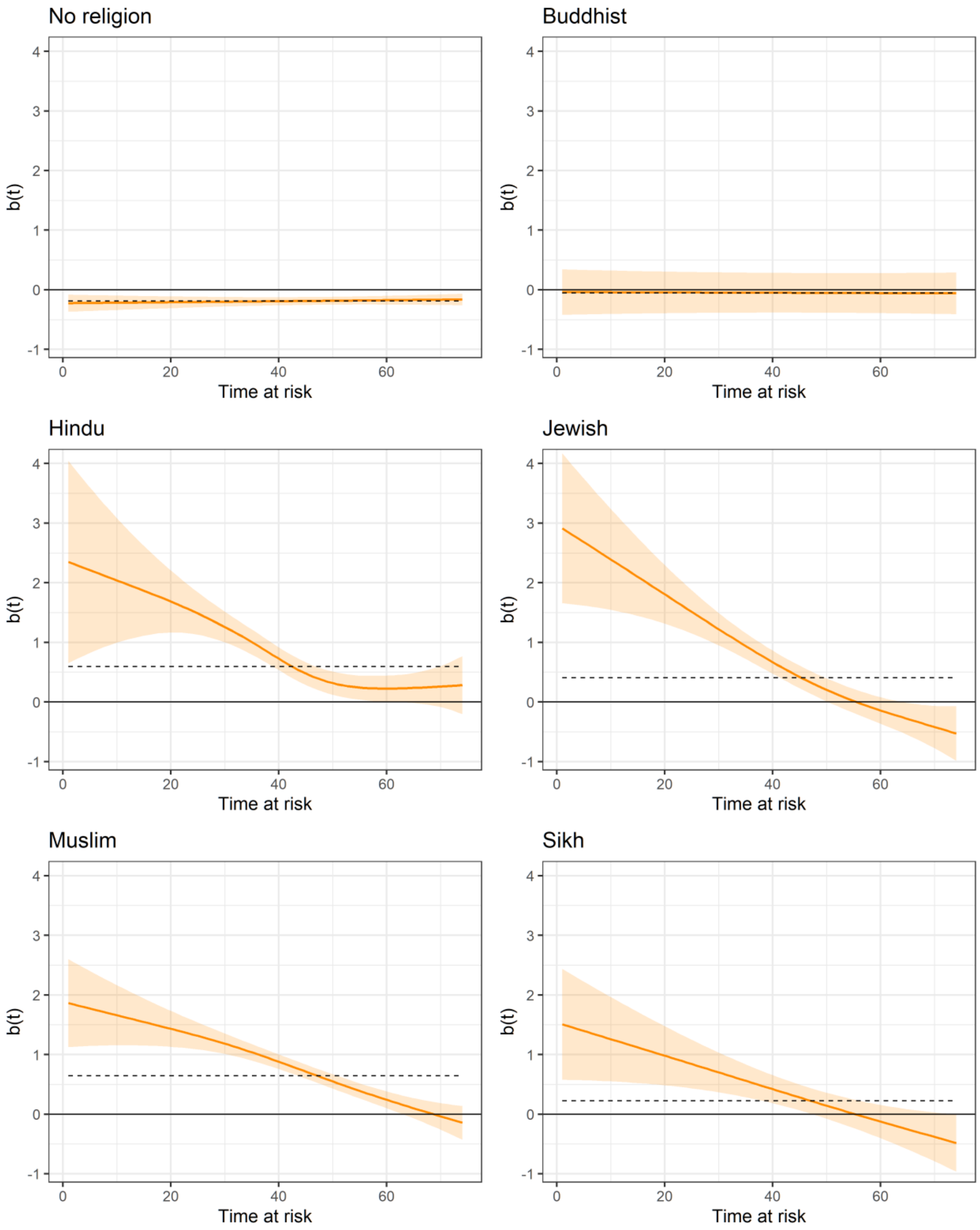

*Supplementary table 3. Pre and post lockdown hazard ratios from extended Cox regression model adjusted for age, males*

| | Pre Lockdown | Post Lockdown |
|---|---|---|
| **No religion** | 0.85 (0.78-0.81) | 0.92 (0.81-0.85) |
| **Buddhist** | 1.46 (0.93-1.09) | 2.29 (1.09-1.49) |
| **Hindu** | 2.59 (2.17-1.63) | 3.08 (1.63-1.86) |
| **Jewish** | 3.53 (2.94-1.63) | 4.25 (1.63-1.92) |
| **Muslim** | 3.97 (3.56-1.96) | 4.42 (1.96-2.15) |
| **Sikh** | 2.23 (1.73-1.30) | 2.88 (1.30-1.58) |
| **Other religion** | 0.93 (0.56-0.89) | 1.55 (0.89-1.21) |
| **Religion not stated** | 0.90 (0.81-0.87) | 1.00 (0.87-0.93) |

*Supplementary table 4. Pre and post lockdown hazard ratio ratios from extended Cox regression model adjusted for age, females*

| | Pre Lockdown | Post Lockdown |
|---|---|---|
| **No religion** | 0.97 (0.85-0.79) | 1.10 (0.79-0.86) |
| **Buddhist** | 1.18 (0.62-0.89) | 2.28 (0.89-1.30) |
| **Hindu** | 3.53 (2.87-1.35) | 4.35 (1.35-1.61) |
| **Jewish** | 3.02 (2.39-1.10) | 3.81 (1.10-1.34) |
| **Muslim** | 3.60 (3.07-1.46) | 4.21 (1.46-1.65) |
| **Sikh** | 1.98 (1.39-1.06) | 2.82 (1.06-1.35) |
| **Other religion** | 1.10 (0.62-0.74) | 1.93 (0.74-1.05) |
| **Religion not stated** | 0.92 (0.81-0.82) | 1.05 (0.82-0.88) |

*Supplementary table 5. Percentage of each religious group falling in one of two socio economic classifications, 2012 and 2019*

| | Percentage of deaths involving COVID-19 among White ethnic group | Percentage of deaths involving COVID-19 among non-White ethnic groups | Percentage identifying as White at the 2011 Census | Percentage identifying as non-White at the 2011 Census |
|---|---|---|---|---|
| **No religion** | 94.00% | 6.00% | 94.30% | 5.70% |
| **Christian** | 94.10% | 5.90% | 93.00% | 7.00% |
| **Buddhist** | 26.60% | 73.40% | 34.40% | 65.60% |
| **Hindu** | 1.40% | 98.60% | 1.10% | 98.90% |
| **Jewish** | 95.60% | 4.40% | 93.60% | 6.40% |
| **Muslim** | 7.80% | 92.20% | 7.50% | 92.50% |
| **Sikh** | 1.60% | 98.40% | 1.50% | 98.50% |
| **Other religion** | 66.30% | 33.70% | 76.50% | 23.50% |
| **Religion not stated** | 90.60% | 9.40% | 87.40% | 12.60% |

*Supplementary table 6. Percentage of each religious group falling in one of two socio economic classifications, 2012 and 2019*

| Managerial and Professional occupations | | | Routine and Manual Occupations | | |
|---|---|---|---|---|---|
| | **2012** | **2019** | | **2012** | **2019** |
| **No Religion** | 26.59% | 28.89% | **No Religion** | 19.59% | 18.04% |
| **Christian** | 22.91% | 24.88% | **Christian** | 19.31% | 16.76% |
| **Buddhist** | 27.80% | 28.50% | **Buddhist** | 23.53% | 21.39% |
| **Hindu** | 29.55% | 32.51% | **Hindu** | 15.92% | 13.67% |
| **Jewish** | 36.86% | 32.33% | **Jewish** | 4.35% | 5.06% |
| **Muslim** | 10.88% | 12.92% | **Muslim** | 14.14% | 12.86% |
| **Sikh** | 20.41% | 25.03% | **Sikh** | 19.09% | 17.22% |
| **Any Other** | 29.61% | 31.03% | **Any Other** | 16.33% | 15.25% |

*Source: Annual Population Survey 2012 and 2019*
